# Topological network properties of resting-state functional connectivity patterns are associated with metal mixture exposure in adolescents

**DOI:** 10.1101/2022.11.14.22282324

**Authors:** Azzurra Invernizzi, Elza Rechtman, Kristie Oluyemi, Stefano Renzetti, Paul Curtin, Elena Colicino, Claudia Ambrosi, Lorella Mascaro, Alessandra Patrono, Daniele Corbo, Giuseppa Cagna, Roberto Gasparotti, Abraham Reichenberg, Cheuk Y. Tang, Donald R. Smith, Donatella Placidi, Roberto G. Lucchini, Robert O. Wright, Megan K. Horton

## Abstract

Adolescent exposure to neurotoxic metals adversely impacts cognitive, motor, and behavioral development. Few studies have addressed the underlying brain mechanisms of these metal-associated developmental outcomes. Furthermore, metal exposure occurs as a mixture, yet previous studies most often consider impacts of each metal individually. In this cross-sectional study, we investigated the relationship between exposure to neurotoxic metals and topological brain metrics (global and local efficiency) in adolescents. In 193 participants (53% females, ages: 15-25 years) enrolled in the Public Health Impact of Metals Exposure (PHIME) study, we measured concentrations of four metals (manganese, lead, copper and chromium) in multiple biological media (blood, urine, hair, and saliva) and acquired resting-state functional magnetic resonance imaging scans. Using graph theory metrics, we computed global and local efficiency (global:GE; local:LE) in 111 brain areas (Harvard Oxford Atlas). We used weighted quantile sum (WQS) regression models to examine association between metal mixtures and each graph metric (GE or LE), adjusted for sex and age. We observed significant negative associations between the metal mixture and GE and LE (β_GE_ = -0.076, 95% CI [-0.122, -0.031]; β_LE_= -0.051, 95% CI [-0.095, -0.006]). Lead and chromium measured in blood contributed most to this association for GE, while chromium measured in hair contributed the most for LE. Our results suggest that exposure to this metal mixture during adolescence reduces the efficiency of integrating information in brain networks at both local and global levels, informing potential neural mechanisms underlying the developmental toxicity of metals. Results further suggest these associations are due to combined joint effects to different metals, rather than to a single metal.

## Introduction

Exposure to neurotoxic metals and their impact on the brain is a growing worldwide health concern [1]. Metals such as lead and manganese have been shown to readily pass the blood-brain barrier and accumulate within various brain areas, where they exert neurotoxic effects [1,2] and are associated with altered neurotransmission, disrupted synaptic structure [1,3–5] and accelerated neurodegeneration [3,6–8]. These among others key features of the above mentioned metals contributed to define the brain as the target organ for exposure [7,9– 11]. Growing research has identified adolescence as a critical window [12] that is vulnerable to environmental exposure including metals [13]. Few studies investigated the neural mechanisms of metal neurotoxicity throughout this extended window of vulnerability. Findings from these studies have linked metal exposure with alterations in regional brain volume [14–19], and brain metabolite concentrations [20–24] during this period. This heightened vulnerability may be due to rapid growth and differentiation of the brain throughout childhood. Neurotoxic exposures during this critical period can also disrupt behavioral, cognitive, and motor development [13–18]. Despite the breadth of research on the developmental effects of childhood metal exposure, the underlying brain mechanisms behind these observed metal-associated outcomes are still not clear.

Over the past decade, increasing use of functional magnetic resonance imaging (fMRI) provides insight into the mechanisms linking metal exposure and alterations in brain functions [19]. In particular, resting-state functional MRI -task-independent assessment of spontaneous fluctuations in blood oxygen level dependent (BOLD) signal from the brain at rest -has emerged as a novel tool in pediatric populations to investigate the intrinsic functional connectivity of the brain. Different from task-based fMRI which requires participants to engage or respond to stimuli [25], in rs-fMRI participants are instructed to simply lay still in the scanner with their eyes closed, while allowing their mind to roam freely (i.e. not focusing their thought on anything in particular). This facilitates research in younger populations, who may have difficulty completing complex tasks in the scanner [25]. Results from rs-fMRI studies have shown a topological organization of the brain in a highly efficient manner with a high level of local clustering, together with long-distance connections [26]. Graph theory analysis of rs-fMRI data characterizes the topological organization of the brain at rest [27] using metrics such as global and local efficiency, which quantify how efficient the brain is at integrating information at global and local levels, respectively [27]. Global efficiency (GE) provides an indication of how efficiently the information is integrated and exchanged between the different regions of the brain [28,29]. In contrast, local efficiency (LE) measures the ability of the brain to perform functionally specialized and segregated processing within a network, requiring densely interconnected regions within modules [28,29]. Previous results have demonstrated the utility to characterize the topological network organization of the brain by using graph metrics based on rs-fMRI and link them with human behavior [30,31], cognition [26,32] and diseases [33,34]. Recent studies have used rs-fMRI to demonstrate intrinsic functional connectivity patterns in a-priori selected brain regions associated with early life exposure to individual metals (i.e. lead, manganese) [21,35,36]. Our data-driven graph theory approach builds on this foundational research by informing potential neural mechanisms underlying the developmental toxicity of metal mixture exposure during adolescence.

To investigate the impact of metal exposure on the brain, it is critical to consider not only single metal exposures but the mixture of co-occurring neurotoxic metals [37]. Historically, studies measure individual chemical concentrations in individual biological media (i.e., blood, urine, etc). These exposure biomarkers are used as surrogates of total exposure from the environment. However, metals distribute unevenly among biological media and therefore, each medium provides complementary information on different biological processes. Recent studies have started to combine information from multiple biomarkers using statistical approaches that resulted in an improved measure of the total body burden and thus improved exposure characterization [17,38,39]. Exposure, defined as metal mixtures, has been observed to more negatively impact neurodevelopment than exposure to a single metal component [14,16,18,40]. Therefore, examining the effects of metal mixtures on brain function is crucial to better understand the real-world impact of metal exposure on cognition and behavior. In this study, we will use an integrated measure of metal mixtures across multiple media, called multi-media biomarker (MMB) [39], to analyze the impact on the brain of each metal across multiple media.

In this study, we investigate how metal exposure impacts brain network properties in adolescents. We use graph theory metrics to quantify how the brain integrates globally (GE) and locally segregates (LE) information and assess associations between these metrics with metal mixture exposure. To define our metal mixtures, we measured concentrations of four metals (lead (Pb), manganese (Mn), chromium (Cr) and copper (Cu)) in four biological media (blood, urine, hair, and saliva) from 193 adolescent participants enrolled in the Public Health Impact of Metals Exposure study (PHIME) study. Then, using weighted quantile sum (WQS) regression, a statistical method commonly used to assess the impact of chemical mixtures on various health outcomes [41], we examined associations between the metal mixture and each graph metric (GE and LE), adjusting for sex and age. This paper contributes to further understanding the impacts of environmental exposures to a mixture of neurotoxic metals in developmental windows like adolescence.

## Materials and Methods

### Participants

The Public Health Impact of Metal Exposure (PHIME) cohort investigates associations between metal exposure from anthropogenic emissions and developmental health outcomes in adolescents and young adults. Details of the study have been described elsewhere [42,43]. Inclusion criteria included: birth in the areas of interest; family residence in Brescia for at least two generations; residence in the study areas since birth. The exclusion criteria were: having a neurological, hepatic, metabolic, endocrine or psychiatric disorder; using medications (in particular with neuro-psychological side effects); having clinically diagnosed motor deficits or cognitive impairment and having visual deficits that are not adequately corrected. Detailed description of this recruitment process and study design can be found in previous publications [43,44]. A convenience based sample of 202 participants (53% female, ages 13-25 years) were selected to participate in a multi-modal magnetic resonance imaging (MRI) study, PHIME-MRI. They completed multimodal MRI scans, neuropsychological tests, including measures of IQ (Wechsler Intelligence Scale for Children, 3rd edition; WISC-III)[46], memory and motor functions. All participants satisfied eligibility criteria for MRI scanning (i.e., metal implants or shrapnel, claustrophobia, no prior history of traumatic brain injury, body mass index (BMI) ≤ 40). Mn, Pb, Cr and Cu were measured in saliva, hair, blood and urine, for each PHIME-MRI participant. Complete exposure data (i.e., all metals in all media for a total of 16 components), MRI and covariates data were available for 193 participants included in this analysis. A total of 193 participants were included in this analysis, 9 were missing at least one biological marker (Figure S1).

Written informed consent was obtained from parents, while participants provided written assent. Study procedures were approved by the Institutional Review Board of the University of California, Santa Cruz and the ethical committees of the University of Brescia, and the Icahn School of Medicine at Mount Sinai.

### Biomarker measures of exposure

Biological samples including venous whole blood, spot urine, saliva and hair were collected from each subject upon enrollment, as described in detail in previous studies [42,47–49]. Complete overview of biomarkers can be found in Supplementary Material (Figure S1) and Table 1. Biological samples were processed and analyzed for metal concentrations using magnetic sector inductively coupled plasma mass spectroscopy (Thermo Element XR ICP-MS), as described elsewhere [42,47–49].

**Table 1.** Metal concentrations (Mn, Pb, Cr and Cu) measured in blood, urine, hair and saliva collected from 193 adolescent participants PHIME-MRI included in the current study.

| Medium* | Metal | % > LOD | LOD mean ± SE | GM | GSD |
| --- | --- | --- | --- | --- | --- |
| Saliva | Lead | 90.6 | 0.05 ± 0.003 | 0.19 | 3.07 |
| | Chromium | 91.7 | 0.13 ± 0.003 | 0.50 | 3.69 |
| | Manganese | 96.4 | 0.08 ± 0.001 | 3.13 | 2.97 |
| | Copper | 97.4 | 0.35 ± 0.025 | 8.63 | 2.35 |
| Blood | Lead | 100 | 0.16 ± 0.003 | 8.84 | 1.56 |
| | Chromium | 62.7 | 0.19 ± 0.008 | 0.34 | 4.54 |
| | Manganese | 100 | 0.49 ± 0.018 | 8.45 | 1.49 |
| | Copper | 100 | 1.09 ± 0.035 | 586.94 | 1.30 |
| Hair | Lead | 100 | 0.003 ± 0.0001 | 0.09 | 2.97 |
| | Chromium | 100 | 0.004 ± 0.0001 | 0.04 | 2.57 |
| | Manganese | 100 | 0.005 ± 0.0003 | 0.06 | 2.58 |
| | Copper | 100 | 0.04 ± 0.002 | 9.96 | 1.62 |
| Urine | Lead | 98.4 | 0.06 ± 0.003 | 0.36 | 2.17 |
| | Chromium | 96.9 | 0.09 ± 0.004 | 0.28 | 3.07 |
| | Manganese | 80.3 | 0.11 ± 0.003 | 0.24 | 3.69 |
| | Copper | 100 | 0.30 ± 0.009 | 6.01 | 1.85 |

**Note:** GM = Geometric mean; GSD = Geometric standard deviation; LOD = limit of detection; SE = standard error of the mean.
\*Metrics used to measure metal concentrations within each medium were: blood and saliva (ng/mL), hair (ug/g), urine (ug/mL)

### MRI and fMRI data acquisition

Magnetic resonance imaging (MRI) and functional MRI (fMRI) data acquisition was performed on a high-resolution 3-Tesla SIEMENS Skyra scanner using a 64-channel phased array head and neck coil, at the Neuroimaging Division of ASST Spetali Civili Hospital of Brescia. For each participant, a high-resolution 3D T1-weighted structural scan was acquired using a MPRAGE sequence (TR =2400 ms, TE= 2.06 ms, TI=230 ms, acquisition matrix=256×256 and 224 sagittal slices with final voxel size=0.9 mm^3^). Fifty contiguous oblique-axial sections were used to cover the whole brain where the first four images were discarded to allow the magnetization to reach equilibrium. For each subject, a single 10-minute continuous functional sequence using a T2*weighted echo-planar imaging (EPI) sequence (TR=1000 ms, TE=27 ms, 70 axial slices, 2.1 mm thickness, matrix size 108×108, covering the brain from vertex to cerebellum) was acquired. During resting-state scans, lights of the MRI room were off, and participants were instructed to stay awake, relax and daydream (not think about anything) while keeping eyes open. They were presented with an image of a night skyline figure projected on a MRI compatible monitor. Padding was used for comfort and reduction of head motion. Earplugs were used to reduce noise.

### fMRI data analyses

Image pre-processing, global and local efficiency calculations, and statistical analyses were performed using SPM12 (Wellcome Department of Imaging Neuroscience, London, UK), Brain Connectivity toolbox [50,51] and customized scripts, implemented in MatLab 2016b (The Mathworks Inc., Natick, Massachusetts) and R (v3.4).

#### Image preprocessing

For each subject, the structural magnetic resonance image was co-registered and normalized against the Montreal Neurological Institute (MNI) template and segmented to obtain white matter (WM), gray matter (GM) and cerebrospinal fluid (CSF) probability maps in the MNI space. FMRI data were spatially realigned, co-registered to the MNI-152 EPI template and subsequently normalized utilizing the segmentation option for EPI images in SPM12. All normalized data were denoised using ICA-AROMA [52]. Additionally, spatial smoothing was applied (8 millimeters) to the fMRI data. No global signal regression was applied.

Based on the Harvard-Oxford [53] atlas, 111 regions of interest (ROI; 48 left and 48 right cortical areas; 7 left and 7 right subcortical regions and 1 brainstem) were defined. For each ROI, a time-series was extracted by averaging across voxels per time point. To facilitate statistical inference, data were “pre-whitened” by removing the estimated autocorrelation structure in a two-step generalized linear model (GLM) procedure [54,55]. In the first step, the raw data were filtered against the 6 motion parameters (3 translations and 3 rotations). Using the resulting residuals, the autocorrelation structures present in the data were estimated using an Auto-Regressive model of order 1 (AR(1)) and then removed from the raw data. Next, the realignment parameters, white matter (WM) and cerebrospinal fluid (CSF) signals were removed as confounders on the whitened data.

#### Graph Theory metrics/Network properties

Global and Local Efficiency (GE and LE) were computed using the Brain Connectivity toolbox [50,51] on the defined ROI time course data per subject. GE and LE build on the concept of efficient integration of communication in a network at local (LE) and whole (GE) level. Based on the average inverse shortest path length in the brain or network, GE is defined as the inverse of the average characteristic path length between all nodes in the networks [56,57]. For each individual node defined as ROI, the shortest number of steps required to go from one node to another was computed. Then, the average number of shortest steps to all defined nodes was computed separately for each node. To correct for the total number of connections between nodes, the inverse of the average number of shortest steps for each node was summed across all network nodes and normalized. LE is computed on the neighborhood of each single ROI/node [50,51] and defined as the inverse of the shortest average path length of all neighbors of nodes among themselves [56]. First, we identified a set of nodes which are directly connected with a given node, then we removed that node from the identified subgraph and calculated the averaged shortest path between all remaining nodes. GE and LE are scaled measures ranging from 0 to 1, with a value of 1 indicating maximum GE/LE in the brain.

### Statistical analyses

#### Descriptive statistics

Visual inspection and descriptive statistics (geometric mean and geometric standard deviation) were used to characterize the metal concentrations in different media. All descriptive statistical analyses were performed using R version 4.2.1.

#### Multi-Media Biomarker (MMB) Approach

To examine associations between our multi-media metal mixture (4 metals, 4 media) and graph theory outcomes (GE and LE), we used a WQS-based multi-media biomarker (MMB) approach [17,58]. Figure 1 shows the complete flowchart of the analysis performed. Briefly, WQS is a data driven, mixtures-based ensemble modeling strategy that tests for associations between the combined effect of multiple, correlated variables and an outcome of interest. The WQS MMB approach builds on WQS, by incorporating exposure information across different biological media, providing an integrated estimate of total bodily exposure to a given chemical as well as identifying the chemical-matrix specific combination that contributes most to the overall association with the graph theory based outcomes (GE and LE) [17]. The MMB WQS is hierarchical with two levels. The first level estimates a weighted index across all biological media for a single metal and the outcome (i.e., Pb MMB = blood Pb, urine Pb, saliva Pb, hair Pb). Our model estimated across 50 bootstrap samples, and 100 repeated holdouts [41] for each individual MMB. By using the repeated holdouts WQS [41], the data are randomly partitioned 100 times to produce a distribution of validated results where the mean is taken as the final estimate. The directionality of the association of the WQS index was constrained to be negative. Note that the WQS assumptions of linearity and directional homogeneity were validated through visual inspection of residuals [17]. The second level estimates a weighted index across the different metals (i.e., Pb MMB, Mn MMB, Cr MMB, Cu MMB) ([17, 59]). First level MMBs are included in the WQS regression model predicting the association between the metal biomarker “mixture” and GE or LE. A significance test for the WQS index provided an estimate of the association with the metal mixture, while the weights associated with each metal MMB provided an indicator of each individual metal contribution to the overall effect. All weights are constrained to sum to one, enabling sorting by relative importance. Metals that impact the outcome have larger weights; factors with little or no impact on the outcome have near-zero weights. These models were adjusted for age and sex, and prior to model estimation, all exposures were grouped into deciles.

**Figure 1.**
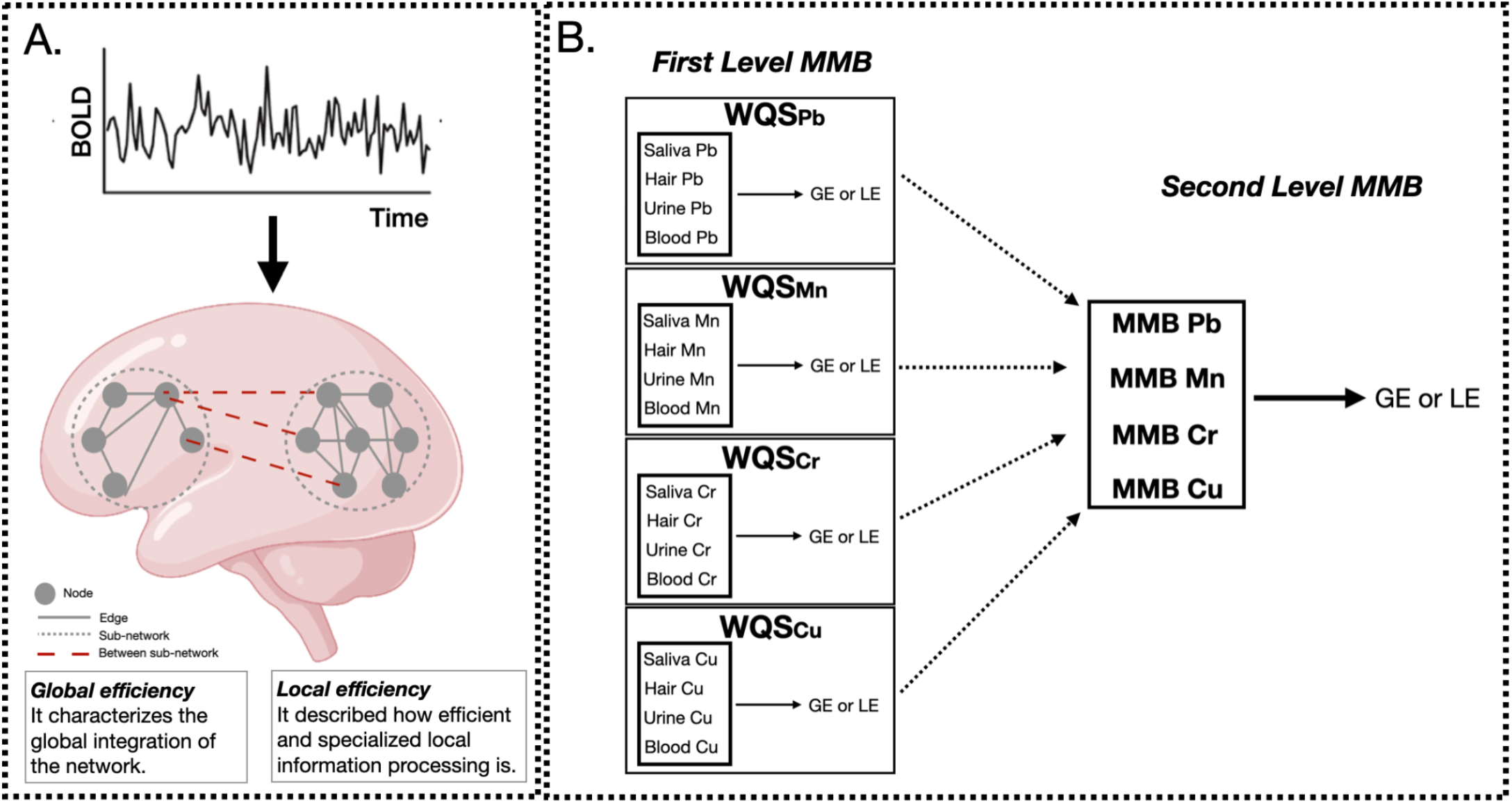
Data analysis flowchart. A.Resting-state fMRI data were preprocessed and the averaged time-series were extracted using the Harvard-Oxford atlas. Then, global and local efficiency (GE and LE, respectively) metrics were computed for each participant using graph theory. Small solid gray circles represent nodes of the graphs (brain regions), while gray connecting lines are the edges of the graph (functional connections). Larger dotted circles represent segregated sub-graphs/networks (functional network characterized by highly connected brain areas), while dashed red lines are the within-network connections at the whole brain level. Panel B shows the two hierarchical levels of analysis performed using the MMB WQS approach to measure the effect of Pb, Mn, Cr, Cu on brain metrics (GE and LE). At the first level, WQS was performed to measure and derive the MMB metric for each metal individually (Pb, Mn, Cr, Cu) on brain metrics (GE and LE). Then, the joint effect of Pb, Mn, Cr, Cu on brain metrics was assessed by applying WQS to Pb, Mn, Cr and Cu MMBs. All models were adjusted for sex and age. Figure adapted from Levin-Schwartz et al. and Rakesh et al.

## Results

### Demographic and exposure characteristics

This study included 193 participants (53% female) living in Northern Italy, with an average age of 19.2 years (range = 15-25) Metal concentrations in the different media are reported in Table 1.

### First level MMBs and brain topological properties

We first examined the association between each individual metal in all media with GE and LE (Figure 2). For all metals, urinary metal concentrations contributed most to the association between the first level MMB (i.e., individual metal in each matrix) and GE. Urinary Pb contributed 46% of the association between Pb exposure and GE. Urinary Mn, Cr and Cu contributed 51, 34 and 68%, respectively to the association with GE. For LE, the most heavily weighted metal-matrix combination differed by metal; blood Pb concentration contributed most to the association with LE (34%). Hair Mn and hair Cr contributed 31% and 43% to the association with LE. Urine Cu contributed the most to the Cu-LE association. Beta coefficients and 95% confidence intervals obtained for each individual MMB WQS model are reported in Figure 2 and supplementary materials (Figure S2).

**Figure 2.**
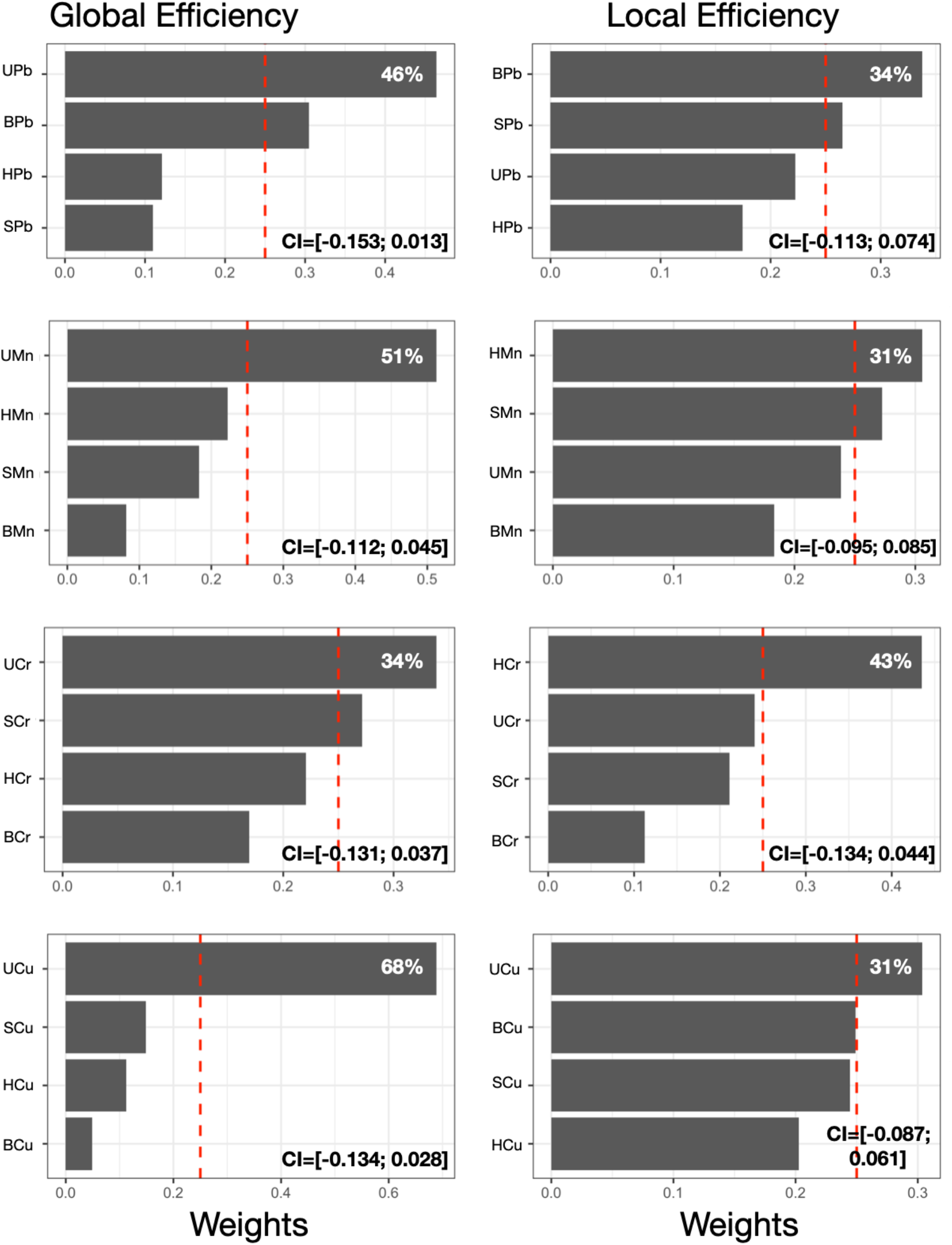
First level MMB models. Results obtained from the MMB WQS association between each metal’s respective exposure biomarker (e.g., blood Pb) and GE or LE was estimated among 193 adolescents included in the current study. Bar plots show estimated weights for each component of the mixture in the WQS regressions. Red dotted lines represent the significant thresholds for each WQS model. Beta coefficients and 95% confidence intervals obtained for each individual MMB WQS model are reported. All models were adjusted for sex and age. Components abbreviations: the first letter represents the medium (S=saliva, B=blood, U=urine, H=hair) and the second and third letters represent the metals (Mn=manganese, Pb=lead, Cr=chromium, Cu=copper).

### Second level MMB and brain topological properties

Results from second level repeated holdout WQS analyses revealed associations between the overall metal mixture and GE and LE (Figure 3a), and the contribution of each metal MMB to these associations (Figure 3). We observed significant negative associations between the combined metal mixture MMB and both GE (β_GE_ = -0.076, 95% CI [-0.122, -0.031]; Figure 3a), and LE (β_LE_= -0.051, 95% CI [-0.095, -0.006]; Figure 3a). We observed that Cr and Pb contributed most to the association between the combined metal mixture and GE (29%; Figure 3b), whereas Cr contributed most to the association with LE (38%; Figure 3c).

**Figure 3.**
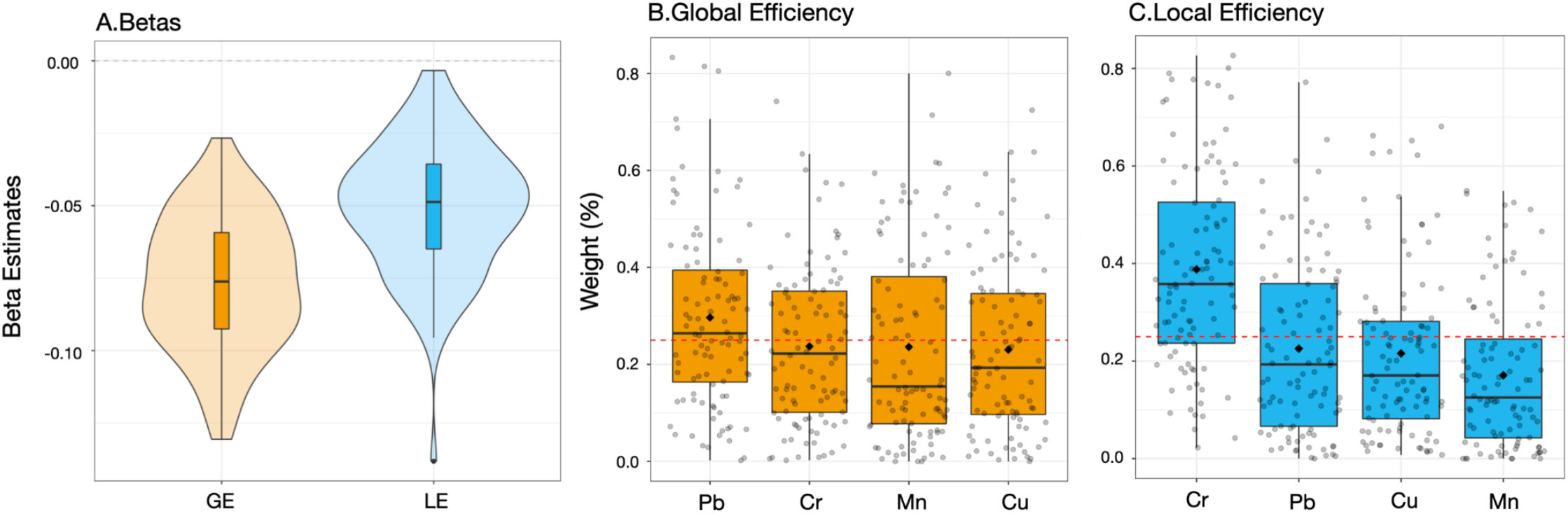
Second level MMBs approach. Beta coefficients (A) and weights (B-C) obtained from the WQS association between each MMB metal and GE, or LE was estimated among 193 adolescents included in the current study. Panel a reports the beta coefficients for GE (orange) and LE (blue), respectively. In panels b and c, data points indicate weights for each of the 100 holdouts; box plots show the 25th, 50th and 75th percentiles while the whiskers show the 10th and 90th percentiles of weights for the 100 holdouts. Diamonds show the mean weights for the 100 holdouts. Dotted lines indicate the thresholds. Mn=manganese, Pb=lead, Cr=chromium, Cu=copper.

## Discussion

This is the first study to use rs-fMRI to investigate global and local connectivity in adolescents exposed to a neurotoxic metal mixture. Using graph-theory based network metrics and a multimedia biomarker (MMB) approach, we observed a significant negative association between exposure to a mixture of five neurotoxic metals (lead, manganese, copper, chromium, and zinc) and global efficiency (GE), with lead and chromium contributing most to this association. Significant negative associations between the metal mixture and both GE and LE were found (β_GE_ = -0.076, 95% CI [-0.122, -0.031]; β_LE_= -0.051, 95% CI [-0.095, -0.006]). We also observed that lead and chromium contributed most to the association with GE (29% and 24%, respectively); while chromium contributed most to the association with LE (38%). Overall, our results substantiate previous findings of associations between metal exposure and altered brain connectivity, and further suggest that environmental exposure to a mixture of neurotoxic metals during adolescence reduces the brain ability to efficiently integrate and segregate information, highlighting the need to further study the impacts of environmental exposures in developmental windows like adolescence [13,60,61]. Furthermore, our results suggest these associations are due to the combined joint effects of multiple metals, rather than to a single metal, emphasizing the importance of analyzing metal mixtures to better understand the real-world impact of metal exposure on brain health.

Our finding that lead and chromium were the top contributing metals in the association between the metal mixture and GE, and chromium contributed most to the association with LE, suggests that these metals may exert a greater influence on global and/or local functional connectivity across/within topological brain networks. Previous human neuroimaging studies have observed associations between lead exposure and altered structural connectivity and functional activation patterns in both children and adults [20–22]. In particular, Thomason et al. found prenatal lead (Pb) exposure was associated with altered age-related intrinsic functional connectivity patterns in developing fetuses [21]. Furthermore, previous studies in animal models have found lead exposure to disrupt multiple neurotransmitter systems [4] (e.g., glutamatergic, dopaminergic, cholinergic), as well as neurotransmitter and synaptic function in various areas of the brain, including the hippocampus [1, 5], and prefrontal cortex [62]. Therefore, our finding of lead being a top contributor to the negative association between the metal mixture and global efficiency, could in part be explained by its impact on structural connectivity (e.g., white matter integrity) and synaptic function and neurotransmission within/across the brain.

While there are no neuroimaging studies investigating the impact of chromium exposure to date, previous studies in humans have observed evidence linking chromium exposure to neurological and psychiatric conditions, including olfactory dysfunction, autism spectrum disorder, and acute schizophrenia [63–66]. These findings suggest an impact of chromium exposure on underlying neurobiological function. Furthermore, previous studies across various animal models have observed brain-wide neurodegeneration following chromium exposure, again suggesting an impact of chromium exposure on neurobiological function via its neurodegenerative effects [63,67,68]. Therefore, our finding of chromium being a top contributor to the negative association between the metal mixture and both local and global efficiency is consistent with these prior studies suggesting its widespread neurodegenerative effects, which could potentially contribute to changes in functional connectivity across brain networks.

Several studies have also detailed the potential synergistic neurotoxic effects of certain metals upon co-exposure, based on their unique chemical properties and similar neurobiological mechanisms of action [69]. Metals within our mixture that have been shown to produce such synergistic neurotoxic effects include lead and manganese [70 -72], whose co-exposure has been observed to increase disruptions to neurodevelopment in both animal [11,17,73,74] and human studies [16,17,75,76]. De Water et al. found that early postnatal manganese (Mn) concentrations were associated with altered intrinsic functional connectivity within cognitive control and motor brain areas of adolescents. Additionally, in another study, de Water et al. found prenatal Mn concentrations were associated with reduced intrinsic functional connectivity of brain areas involved in emotion processing and regulation in children. Furthermore, co-exposures of certain metals have been reported to potentially increase accumulation, retention, and distribution of individual metal components in animal models [72]. In particular, manganese has been shown to increase accumulation of various metals in the brain, notably lead [11,72], and copper [77]. Therefore, while lead and chromium were found to contribute most to the association between the metal mixture and GE, and chromium contributed most to the association with LE, the higher influence of these metals may be due to synergistic interactions with other metals in the mixture (e.g., manganese). This possibility highlights the importance of analyzing metal mixtures rather than single metals in environmental epidemiological studies, as the influence of a single metal exposure may be affected by other metals an individual is exposed to. Further, our findings show that urine contributed most to the association between both lead, chromium and GE, and hair contributed most to the association between chromium and LE, suggesting that urine and hair may be critical biomarkers for estimating the effects of metal mixtures on brain connectivity. Previous studies that analyzed one metal at the time, have indicated blood as the most reliable biomarker to assess lead exposure [17], and blood has also been used previously as an exposure biomarker for other metals such as chromium [63,78] and manganese [17]. By using novel techniques like MMB WQS, we can increase the accuracy in measuring mixture effects compared to individual biomarkers and provide a data-driven biomarker selection [39,59]. Finally, as previous neuroimaging studies have mainly examined associations between brain function and a single metal exposure, future studies should aim to utilize metal mixtures to better account for these potential synergistic effects due to metal co-exposure, which would ultimately help better understand the real-world impact of metal exposure on the brain.

### Limitations

In this study, while we found robust associations between metals and GE and LE metrics, our small sample size resulted in relatively small effect sizes (Figure 3). While it would be beneficial to repeat our analysis in a larger dataset, to our knowledge no such dataset with multi-media biomarkers and fMRI data exists. Further, we assumed that all metals have a linear association with both global and local efficiency metrics. Our MMB WQS approach does not assume linear associations between outcomes but only considers additive effects. Future studies should investigate non-linear associations between outcomes and possible multiplicative effects. Finally, MMB WQS might suffer from overfitting issues, since two WQS models are performed on the same set of data. To compensate for this, we split our data into training and testing datasets in both MMB WQS analysis levels.

## Conclusion

Using a multimedia biomarker (MMB) approach, we were able to estimate the top contributing biomarkers to the associations between individual metals in the metal mixture and GE and LE using the weights associated with each metal MMB. Our finding that urine contributed most to the associations between both lead and chromium and GE, and hair contributed most to the associations between chromium and LE, suggests that urine and hair may be critical biomarkers for estimating the effects of metal mixtures on brain connectivity. Given our results, we suggest that future neuroimaging studies on metal mixture exposure aim to collect urine and hair specimens to explore the effects of metal mixtures on the brain. Altogether, our research supports the notion of adolescence being a timepoint of vulnerability to environmental exposures. More specifically, our results suggest that the adolescent brain connectivity is vulnerable to metal mixture exposures during this period. Given that adolescence is a period of rapid brain development, especially in prefrontal areas, our results suggest that metal exposure may have the potential to alter neurodevelopment via changes to global and local connectivity. These connectivity changes may potentially lead to alterations in cognition and neurobehavior in adolescence. Therefore, future environmental neuroimaging studies should focus on adolescents to further characterize how metal mixture exposure during this period can lead to potential alterations in brain development (e.g., brain volume, functional connectivity), and ultimately neurobehavior and cognition.

## Data Availability

All data produced in the present study are available upon reasonable request to the authors

## Acknowledgements

The authors would like to acknowledge support from the National Institutes of Environmental Health Sciences (NIEHS) grants numbers R01 ES019222, R01 ES013744, P30ES023515, and the European Union through its Sixth Framework Programme for RTD (contract number FOOD-CT-2006-016253).

## Supplementary Materials

**Figure S1.**
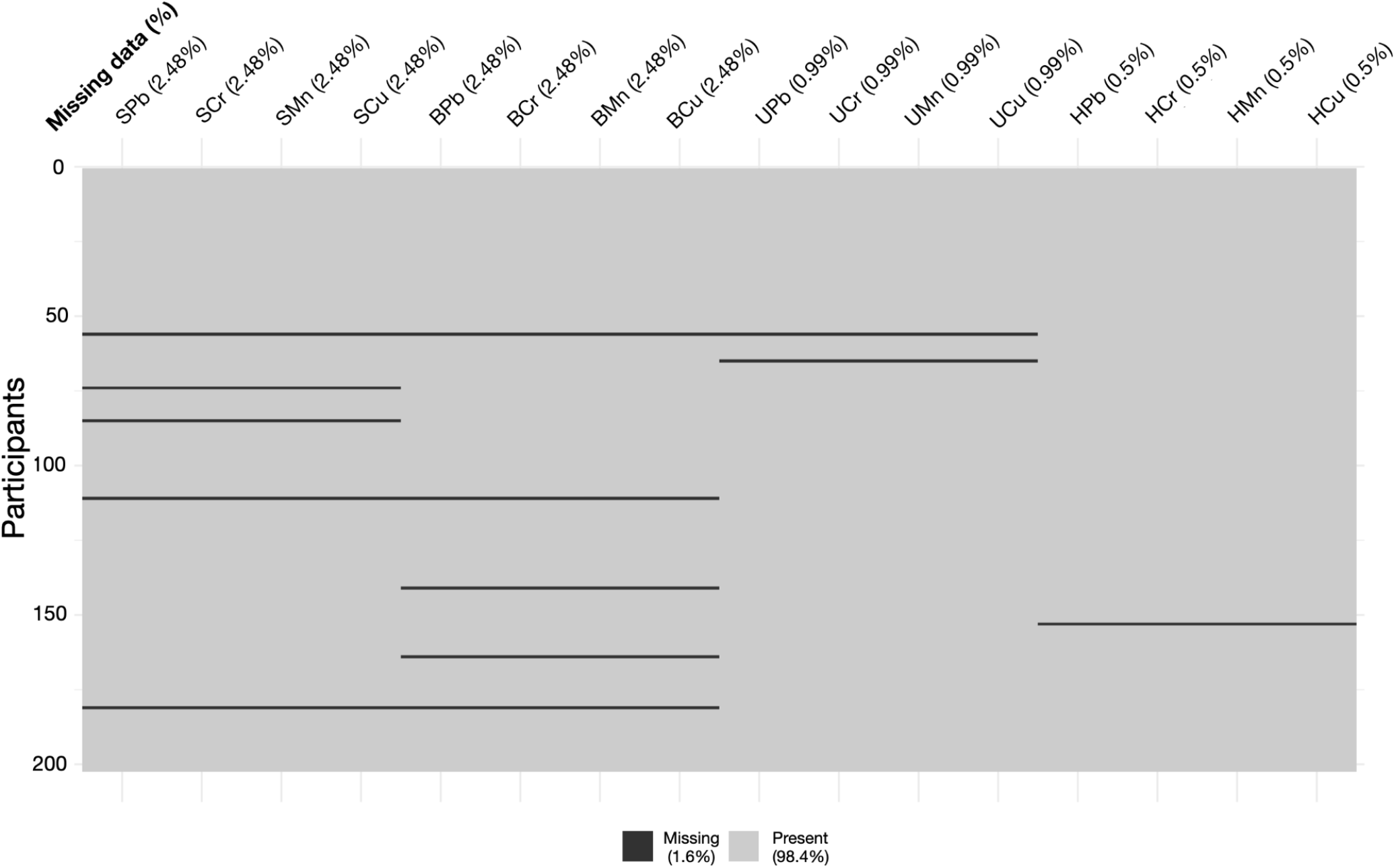
PHIME-MRI biomarker. Complete overview of all biomarkers collected in PHIME-MRI. The dark lines represent the missing values while the gray ones the collected and analyzed values for each PHIME-MRI participant. Percentage of missing data for each component is reported on the top of the figure, after the component’s names. Components abbreviations: the first letter represents the medium (S=saliva, B=blood, U=urine, H=hair) and the second and third letters represent the metals (Mn=manganese, Pb=lead, Cr=chromium, Cu=copper).

**Figure S2.**
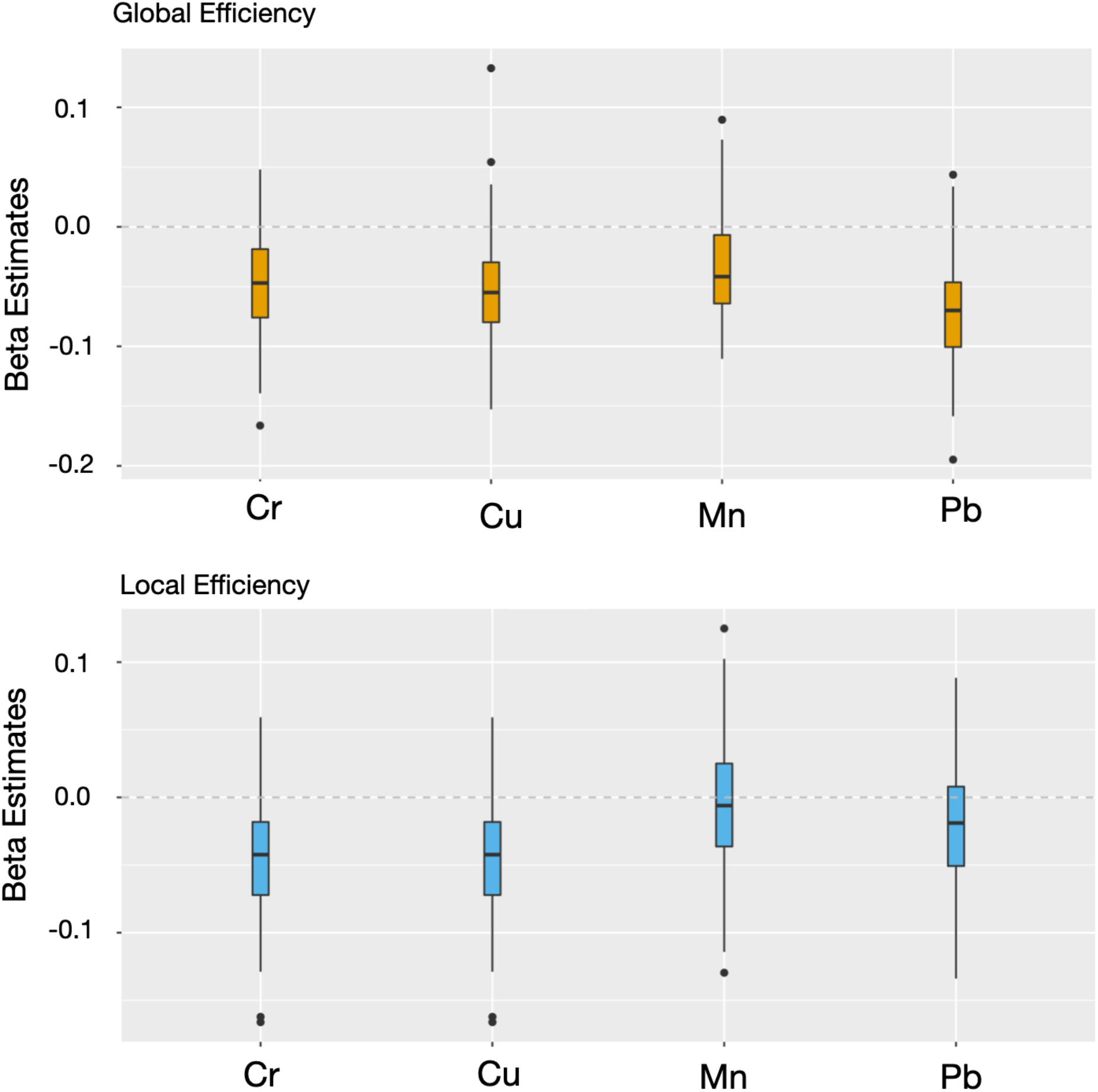
MMBs approach: first level. Betas and 95% confidence interval obtained from the WQS association between each metal’s respective exposure biomarker (e.g. blood Pb) and GE (orange bar) or LE (blue bar)was estimated among 192 adolescents included in the current study. All models were adjusted for sex and age. Components abbreviations: the first letter represents the medium (S=saliva, B=blood, U=urine, H=hair) and the second and third letters represent the metals (Mn=manganese, Pb=lead, Cr=chromium, Cu=copper).

